# Improved morpho-syntax in discourse following intensive voice treatment in Parkinson’s disease: Secondary outcome variables from a Randomized Controlled Trial (RCT)

**DOI:** 10.1101/2021.10.07.21263659

**Authors:** Amy E. Ramage, Kathryn J. Greenslade, Kaila Cote, Jessica N. Lee, Cynthia M. Fox, Angela Halpern, Lorraine O. Ramig

## Abstract

It is well established that voice is disordered in nearly 90% of individuals with Parkinson’s disease (PD). Given the role of voice in language expression, we pose that optimizing vocal function may lead to improved language production. Verb production is an area of language deficit in PD, particularly for verbs associated with an individual’s location of impairment (upper vs. lower limbs). It is thought that damage to the motor system, given its connection to action verbs, underlies this lexical effect. If this is the case, then treatment improving vocal motor function may also improve access to verbs. Nineteen participants with PD underwent Lee Silverman Voice Treatment (LSVT LOUD®), a 4-week intensive voice treatment (TXPD), in an IRB-approved randomized controlled voice treatment trial. Language production was contrasted with 20 untreated PD (UNTXPD) and 20 age-matched neurotypical control participants. Each provided 1-minute picture description narratives at baseline and after 4-weeks. Pre-post treatment within- and between-group comparisons identified effects of assessment time point and isolated treatment effects in the TXPD relative to UNTXPD and Controls. Given the intervention, the TXPD group demonstrated a significant increase in loudness during the picture description, as well as increased utterance length, diversity of word types used, verbs per utterance, and lexical density.

## 1. Introduction

Language and action are intertwined. This is true from the point of word and rule acquisition to the moment-to-moment formulation of utterances. According to the embodied cognition theory, the cognitive systems that represent body, motor, and sensory perception are integral to language processing (Wang et al., 2019). Voice and speech, as components of the motor system, are the vehicles on which oral language is expressed. It stands to reason, then, that the neural systems underlying movement and expressive language production must be intertwined. However, motor and language systems are frequently considered separately, particularly when approaching remediation of either.

An example of this is in individuals with Parkinson’s disease (PD) who have characteristic impairments in vocal motor control secondary to dopaminergic deficiencies (Berardelli et al., 2001; C. M. Fox et al., 2002; Hallett & Khoshbin, 1980). PD results in hypokinetic dysarthria in up to 89% of patients (Hartelius & Svensson, 1994; Ho et al., 1998; Logemann et al., 1978; Miller et al., 2007; Schalling et al., 2017), affecting vocal quality (i.e., reduced loudness, hoarse and/or breathy quality, and monotone prosody; (Baumgartner et al., 2001; C. M. Fox & Ramig, 1997; Ho et al., 1999; Midi et al., 2008; Plowman-Prine et al., 2009) as well as speech clarity (i.e., articulatory precision, vowel centralization) and speech rate (Ackermann & Ziegler, 1991; Forrest et al., 1989; Logemann & Fisher, 1981; Sapir et al., 2007, 2010; Skodda & Schlegel, 2008; Tjaden et al., 2013; Tjaden & Wilding, 2004).

Language is also altered in individuals with PD. More specifically, action word or verb processing is thought to be diminished (Johari et al., 2019; Péran et al., 2009; Rodríguez-Ferreiro et al., 2014; Salmazo-Silva et al., 2017), particularly for verbs associated with an individual’s location of greatest impairment (upper vs. lower limbs; (Roberts et al., 2017)). As well, access to or production of highly actionable nouns (e.g., screwdriver, toothbrush) is represented across semantic and motor domains (Noppeney et al., 2006), but is minimally impaired in PD, if at all (Bocanegra et al., 2017). These deficits in action word processing tend to be verb-specific, are seen relatively early in PD pathogenesis, and are evidenced in access to lexical representations for the comprehension and production of actions, both in isolation and in contextual interactions (e.g., (Cardona et al., 2013)). However, verb processing in individuals with PD is not necessarily associated with motor impairment severity, unlike in other motor disorders (e.g., amyotrophic lateral sclerosis [ALS] in (Cousins et al., 2018); cervical dystonia in (Bayram & Akbostanci, 2018)). These findings indicate a unique interaction between the motor and language systems in PD, likely centered in cortico-striatal loops (Cardona et al., 2013; Silveri et al., 2012). Several investigations of action word processing in individuals with PD or with other motor system disorders have isolated the representations of action concepts to the central neuromotor systems, rather than in peripheral motor systems (Cardona et al., 2014). This suggests that action verbs may have inextricably intertwined representations in the motor and language systems. More specifically, investigations of several motor disorders have parsimoniously localized aspects of action concepts to cortico-striatal loops (as impaired in individuals with PD), rather than to primary motor or premotor cortex (as is impaired in individuals with ALS, (Cousins et al., 2018)). The latter finding highlights the potential for effector-specific representations of the semantics for action concepts, a hypothesis that is best couched in the theories of embodied cognition (Barsalou, 2010, 2020; Binder & Desai, 2011; Pulvermüller, 2005).

What remains unclear is whether action word processing in PD is attributable to a motor system impairment influencing language processing specifically, or if it is related to the presence of mild cognitive impairments that may be present in this population (c.f., (Bocanegra et al., 2017; Murray, 2000)). Subtle cognitive differences are observed in PD, particularly in executive functioning and processing speed (Schapira et al., 2017; Smith & Caplan, 2018), which could explain impaired language in contexts of higher processing load like discourse (e.g., increasing syntactic complexity, trace movement distance). Thus, the differences observed in verbs for PD may be attributable more generally to the cognitive deficits influencing sentence processing and the need for executive planning and working memory demand for more syntactically complex stimuli. There also is evidence suggesting that motor system modulation alone affects linguistic processing in PD, particularly for action words.

One obvious approach to investigating the motor system in PD is through the changes observed in performance when individuals are on or off dopaminergic medications. Individuals with PD who are taking dopaminergic medications produce more action verbs and action content in a verbal fluency task compared to those who are not taking medications (Herrera et al., 2012). Herrera and colleagues concluded that “the dopamine network from basal ganglia to brain motor areas might play a role in retrieving action verbs with specific semantic representations” (p. 72). This would suggest that the deficit is at the early stages of message formulation, at the level of lexical semantics (F. Ferreira, 2000) but is also likely influenced by the task employed (e.g., verbal fluency in the Herrera study, for which lexical retrieval is the target behavior).

A similar approach to investigating dopaminergic influence on language has been to investigate individuals with PD who have undergone surgical implantation of deep brain stimulators. A small number of studies of language performance on story generation or spontaneous speech samples assessed pre- and post-surgical stimulator placement in the subthalamic nucleus (STN, (Zanini et al., 2003)) or the pedunculopontine nucleus (PPN, (Zanini et al., 2009)) have reported improvements in morphosyntax with stimulator on versus off. That is, the structure of words (morphology) and the grammar (syntax), both of which are thought to be guided by verb selection (c.f., (F. Ferreira, 2000)), improved with stimulation. Despite the small sample sizes, no changes (good or bad) were observed pre- to post-surgery or on/off stimulation for neuropsychological measures or lexical semantics (words and meanings), suggesting specificity of effects in discourse production at the later stages of utterance planning – i.e., at surface structure for grammatical encoding, function assignment, or constituent assembly (Bock & Levelt, 1994; V. S. Ferreira, 2010).

These findings support, in part, the embodied cognition theory, which posits that language production, particularly that associated with action/verb processing, may vary with modulation of the motor system in individuals with dopaminergic degeneration. A logical extension of this theory is that improving motor system function may improve language function. Thus, the causal hypothesis of the present study is that if motor system dysfunction impairs action word processing, then treatment improving motor system function should also improve access to action words. In turn, if action word access is improved, then expressive morphosyntax may improve given the central role of the verb in sentence formulation (F. Ferreira, 2000).

The current study aims to clarify the interconnectedness of the motor and language systems by determining whether treatment targeting improved motor function in PD is associated with improvements in language production. PD is an optimal population for investigating whether remediation of one effector system may influence the other given that the disease is known to affect the nigrostriatal pathways of the motor system and, in many cases, does not involve more diffuse cognitive deficits early in the pathogenesis of the disease (c.f., Schapira et al., 2017).

For this study, secondary analysis was conducted on data from a randomized controlled trial of Lee Silverman Voice Treatment (LSVT LOUD®). This intervention has been shown to effectively improve vocal loudness in individuals with PD (Ramig et al., 2018) as well as improvements in other areas of speech motor function and communication including speech articulation (Sapir et al., 2007), intonation (Ramig et al., 2001), speech intelligibility (Levy et al., 2020; Schulz et al., 2021), facial expression (Dumer et al., 2014), swallowing (El Sharkawi et al., 2002; Miles et al., 2017), and neural functioning related to speech (Baumann et al., 2018; Narayana et al., 2010). This treatment targets change at the level of motor function (respiratory-laryngeal-oromotor support), which may affect system-wide improvements including changes in language production. Specifically, language production in a narrative discourse task was investigated prior to and immediately following this intervention. The aims of the study are to (1) replicate differences in discourse-level language production and information content in individuals with PD relative to neurotypical, age-matched healthy controls, and (2) determine whether participation in LSVT LOUD leads to improvements in discourse-level language production or information content. We hypothesize that differences in discourse-level language production will be observed between individuals with PD and controls at baseline. Further, we hypothesize that LSVT LOUD will yield improvements in discourse-level language. If such improvements are specific to verb production – either in terms of retrieval of verbs (lexical semantics) or in the use of verbs in context (morphosyntax), this result would lend support to the theory of embodied cognition. Finally, as a preliminary investigation into whether motor system function is associated with changes in language production following intervention, correlations between pre- to post-intervention change in motor performance (e.g., changes in vocal sound pressure levels/loudness) and change in language production are computed. We hypothesize that changes in the motor system will lead to system-wide changes that will be associated with changes in language production following treatment.

## 2. Material and methods

### 2.1 Participants

Participants in the present study are a sub-set of those reported in the LSVT LOUD protocol of Ramig et al. (2018). Participants with PD were diagnosed by a neurologist, were between stages I to IV on the Hoehn and Yahr scale for symptom severity (Hoehn & Yahr, 1967) and were stable on antiparkinsonian medications. All were 45-85 years of age with normal hearing for age, scored ≥ 25 on the *Mini-Mental Status Examination* (MMSE, (Folstein et al., 1975)), and reported no greater than moderate symptoms of depression (based on the *Beck Depression Inventory II* ≤ 24, (Beck et al., 1996). Individuals were excluded if they had PD with other neurological conditions or atypical PD at the time of screening (e.g., multi-system atrophy, palilalia), presented with speech or voice disorders unrelated to PD, had undergone neurosurgical treatment, had laryngeal pathology or surgical history, or had swallowing impairment requiring treatment were excluded. Additionally, individuals with PD who had undergone intensive speech treatment within the 2 years prior to the study or who had participated in LSVT LOUD previously were excluded.

Study procedures were approved by the institutional review board at the University of Colorado at Boulder and are shared at ClinicalTrials.gov identifier: NCT00123084. All participants provided informed written consent for participation. Please see Ramig et al. (2018) for details regarding screening and randomization for the LSVT LOUD treatment trial.

Twenty participants with idiopathic PD underwent LSVT LOUD (TXPD), targeting the respiratory-laryngeal-oromotor systems, for the full 4-week intensive dose. Participants received 16, one-hour sessions of voice treatment, delivered four days a week over four weeks. The first 30 minutes of each treatment session targeted: (*a*) a minimum of 15 maximum duration sustained vowels (e.g., “say ‘ah’ for as long as you can using your loud voice”); (*b*) a minimum of 15 maximum f_0_ range (e.g., “say ‘ah’ as high/low as you can with your loud or big voice”); and (*c*) a minimum of 5 repetitions of 10 functional phrases selected by participants as phrases used in their daily communication. The second 30 minutes of each treatment session included individualized speech hierarchies designed to maximize functional communication goals. Speech hierarchies progressed both in length of utterance (words, phrases, sentences, etc.) and complexity (repetition, reading, conversation, communication activities, with/without distractors). Further, for each participant, speech hierarchy practice materials were made meaningful to each participant by selecting topics of interest, hobbies or activities related to functional communication goals for each participant. For example, in week 1 of treatment, the tasks might include structured reading at the word/phrase level and spontaneous single word/phrase level responses to questions, word association tasks, fill-in-the-blank or simple descriptions. In week 2, the target would increase to structured reading at the sentence level and spontaneous short/simple conversation. Week 3 might involve structured reading or spontaneous conversation at the paragraph level; week 4 might target continuous conversation. During all these tasks, participants were required to maintain their target loudness giving maximum effort. Further, for each participant, speech hierarchy practice materials were made meaningful by selecting topics of interest, hobbies, or activities related to functional communication goals. Throughout the entire one-hour session, focus on sensory awareness and functional goals were emphasized to encourage generalization (e.g., “Feel that effort when you use your loud voice? That is what you need to feel when you answer the phone at work.”). All participants were required to do homework once per day on the day of treatment and twice per day on days when treatment did not occur. Homework involved repetitions of the same three exercises from the first 30 minutes of treatment sessions, as well as hierarchy practice and carry over assignments (i.e., ordering dinner at a restaurant using the target louder voice) (see Fox et al., 2012 and Ramig et al., 2018 for the full protocol).

To ensure treatment fidelity, three LSVT LOUD certified speech-language pathologists administered treatment following established protocols, including instructions about encouragement and positive reinforcement during treatment, ensuring treatment fidelity. Additional information about the training of clinicians, control of bias, and maintaining treatment fidelity are presented in Levy et al.(2020) and Ramig et al. (2018).

A second group of 20 participants with idiopathic PD served as an untreated comparison PD group (UNTXPD), and 20 age-matched, neurotypical participants served as healthy controls (Controls). All experimental measures were acquired from participants at baseline (baseline, pre-intervention in the TXPD group) and after 4-weeks (one-month, immediately post-intervention in the TXPD group).

### 2.2 Picture Description and dB SPL Data Acquisition

Participants were assessed on a range of speech tasks during the data collection session, including a description of the Cookie Theft Picture (*Boston Diagnostic Aphasia Examination;* (Goodglass et al., 2001), which is the focus of this study. For the picture description task, participants were asked to describe the Cookie Theft picture in as much detail as they could for about a minute. To measure vocal loudness during this task, audio recordings were obtained in a sound-treated booth with the participant seated in a dental or straight-back chair using a head mounted microphone placed at eight centimeters from the mouth. The microphone was calibrated to a Type I Sound Level Meter to extract decibels (dB) of SPL. The microphone files were edited to eliminate coughs, extraneous talking not related to the task, etc. The edited, calibrated microphone files were analyzed for SPL resulting in a mean and standard deviation value for dB SPL at a reference distance of 30 cm.

### 2.3 Transcription, Segmentation, and Coding Procedures

Audio recorded language samples were transcribed using CHAT conventions in the CLAN program (V 30-Jan-2020, (MacWhinney, 2000)). Transcripts were separated into conversational units, or C-units, defined as a main clause (noun + verb) and all dependent clauses attached (Loban, 1976). Rules for determining the end of an utterance included the participant using: (1) the coordinating conjunctions (to connect two main clauses): *for*, *and*, *nor*, *but*, *or*, *yet*, or *so*; (2) terminal intonation contour; or (3) complete grammatical structure (the main clause and all dependent clauses attached).

Two graduate student transcribers (KC and JL), naïve to participant group, time point, or any other identifying information, were trained with six examples to establish reliability. They then independently transcribed 10 samples each. Once interrater reliability was sufficiently established (>.80 for intraclass correlation coefficient (ICC) and point-to-point), the remaining 114 samples were transcribed in four blocks to monitor reliability and prevent drift. Then, EVAL (MacWhinney, 2000) was run for each transcript, outputting measures and scores for various language features. Programs including FLUCALC and frequency of verbs were also run for fluency and verb analysis, respectively (MacWhinney, 2000). The accuracy of verbs coded by CLAN was checked (KC). Due to an error rate in verb coding of 7.46% (e.g., “socks” coded as a verb), three graduate students (KC, JL, and a third research assistant) examined all verbs to determine whether they were coded correctly with consensus across coders and re-coded as needed to reflect the correct part of speech.

Based on analyses of transcription and coding, the following dependent variables were utilized:

1. Lexical semantic variables (related to the retrieval of words, especially verbs): a) the number of different words (types), b) the total number of words (tokens), c) the ratio of types and tokens (type-token ratio; TTR), which is an index of lexical diversity and d) the percentage of verbs and nouns to the total number of words.
2. Morphosyntactic variables (addressing the inclusion of verbs to form complete utterances and the syntactic complexity of those utterances): a) the total number of utterances, b) the mean length of utterance (MLU) in words or morphemes, c) the number of verbs per utterance, and d) the lexical density of propositional ideas, which is approximated by the number of verbs, adjectives, adverbs, prepositions, and conjunctions to the total number of words (MacWhinney, 2000).

Thirty-four language samples (29.82%) were checked for reliability. For each sample, a two-way random ICC with absolute agreement was computed using SPSS Statistics software (version 26.0.0.0, (IBM Corporation, 2015)). In addition, point-to-point reliability for utterances (91.59%) and words (94.55%) was calculated for thirty samples. Disagreements were discussed and resolved by consensus. As summarized in Supplemental Table 1, interrater reliability for the variables derived from EVAL and FLUCALC ranged from moderate (ICC(2,1) = .50-.75) to excellent (ICC(2,1) > .90).

### 2.4 Discourse Information Analysis Coding Procedures

Main concepts and content units were coded manually by the same two graduate students who completed transcriptions. Main concepts were coded for the presence, accuracy, and completeness of seven statements that define the essential elements for the Cookie Theft picture (Nicholas & Brookshire, 1995). Accurate and complete productions had to include each element of the main concept, allowing for alternative productions if they conveyed the same meaning. For example, the statement, “the little **boy** that’s up **on the stool** that’s about ready **to fall**,” was considered accurate and complete for two main concepts: #3 “the ^1^**boy** is ^2^**on** a ^3^**stool**” and #5 “the ^1^**stool** is ^2^**tipping**” (where numbered/bolded words indicate essential elements). As illustrated in this example as well as in additional examples from Appendix B of Nicholas and Brookshire (1995, p. 155-156), utterances that contained two main concepts were broken up according to the following rules: 1) each main concept should have a subject, verb, and all necessary objects and 2) if the utterance contains only one subject but two verbs separated by “and,” the subject may be reused. At times, prepositional phrases were divided from utterances with a single subject and verb to allow participants to receive partial credit for more than one main concept. If a participant produced more than one utterance that could be matched with a single main concept, the utterance that was the most accurate and complete was coded (as opposed to coding the final version as originally suggested by Nicholas & Brookshire, 1995). For each main concept that was present, the participant’s matching utterance was coded as accurate and complete (AC, 3 points), accurate but incomplete (AI, 2 points), inaccurate but complete (IC, 2 points), or inaccurate and incomplete (II, 1 point). Absent main concepts (AB) received a score of 0 points.

In addition to main concept analysis, Yorkston and Beukelman’s (1980) 56 content units for the Cookie Theft picture were counted for each participant, accounting for alternative wordings. In this coding system, if participants mentioned a content unit twice, only one use was counted. Additionally, if a participant’s utterance matched two content units, only one was counted.

Each of the transcribers/coders completed three training examples to learn main concept analysis and coding of content units. Disagreements were discussed and resolved with the lead researchers (AER and KJG). For main concept analysis, reliability training continued until interrater reliability was deemed adequate (kappa > 0.75, point-to-point > 0.8) for at least 80% of transcripts in a set (12 of 15 transcripts met this criterion in the final set; note that these 15 samples were not counted toward the reliability sample, as they contributed to training). For content units, reliability was excellent (kappa and point-to-point > 0.9) for all training examples.

Once interrater reliability was established, the transcribers/coders proceeded with coding of main concepts and content units in four blocks to monitor reliability and prevent drift. For main concepts, 24 of 99 samples (24.2%) were checked for reliability, yielding an average *k*=.809 and point-to-point reliability of 82.2%. For content units, 27 of 114 samples (23.7%) were checked for reliability, yielding an average *k*=.929 and point-to-point reliability of 92.9%. Consensus data were created following reliability analyses and used for study analyses.

### 2.5 Data Analysis

Because the current study represents secondary analyses of an RCT, the original study was powered for the planned analyses of the original study. However, the current study excluded four of the original study participants due to atypical PD symptoms (2 TXPD and 1 UNTXPD excluded for multi-system atrophy, 1 UNTXPD excluded for presence of palilalia). The outcomes for the present study included 9 dependent variables (words per minute, word types and tokens, type-token ratio, number of utterances, MLU words, MLU morphemes, verbs per utterance, and density) while the original study had one (vocal loudness in dB SPL). The G*Power 3.1.9.4 software package was used for re-calculation of power (Faul et al., 2007) including the 9 dependent variables and 3 groups, indicating that the total sample size of n = 60 provided 84% power to detect a large effect size (*f^2^*= .35) and 60% power to detect a medium effect size (*f^2^*= .15) for assessing group differences at baseline and for differences between assessment timepoints by group (repeated measures, within-between interaction).

#### Between-group Contrasts for Baseline

All variables were assessed for outliers and for normality of distribution (Shapiro-Wilk test of normality) in SPSS. Several of the language variables were not normally distributed, and there was considerable variance within groups (Table 2). Thus, generalized mixed models were used to analyze group differences at baseline and for change in performance between baseline and one-month assessments. These models allow for the accommodation of different distributions for the dependent variables as well as the inclusion of random and residual effects. Comparisons of model fit (specifically the Akaike corrected information criterion) elucidated whether inclusion of random effects (participant, residual variance) improved the models. *P* values < .05 were considered significant and post-hoc pairwise comparisons were controlled for multiple comparisons with the Bonferroni method.

**Table 1.** Subject demographics and symptoms. The groups did not differ for age (*F*(2,56) = 0.48, *p* = .62), or for gender (*X^2^_4_* = 1.18, *p* = 0.88) or handedness (*X^2^_4_* = 5.96, *p* = 0.65) distribution, or for *Mini-Mental Status Exam* (Folstein et al., 1975) score (*H*(2) = 2.23, *p* = 0.33). The TXPD group endorsed more symptoms of depression on the Beck Depression Inventory II (BDI, Beck et al., 1996) (*H*(2) = 17.32, *p* < .0001), with three participants scored above the clinical cutoff of 17 (all in the TXPD group – IDS74 = 18; IDS06 = 19, IDS05 = 20). The PD groups did not differ for motor symptom severity (Hoehn & Yahr scale, (Bhidayasiri & Tarsy, 2012) (*U*(2) = 165, p = 0.50)) or years since diagnosis (*U*(20) = 170.5, *p* = 0.59).

|  | TXPD | UNTXPD | Control |
| --- | --- | --- | --- |
| **n** | 19 | 20 | 20 |
| **Age** | 67 ± 8 | 65 ± 9 | 64 ± 9 |
| **Gender (M:F)** | 15:4 | 13:7 | 13:7 |
| **Handedness (R:L)** | 17:1 | 16:4 | 19:1 |
| **Beck Depression Inventory II** | 9.47 ± 6 | 7.3 ± 5 | 2.85 ± 3 |
| **Mini-Mental Status Exam** | 28.8 ± 1 | 29.0 ± 1 | 29.3 ± 1 |
| **Hoehn & Yahr Scale** | 2 ± 0.8 | 2.03 ± 1 | - |
| **Years Since Diagnosis** | 4.83 ± 7 | 4.73 ± 4 | - |

**Table 2.**
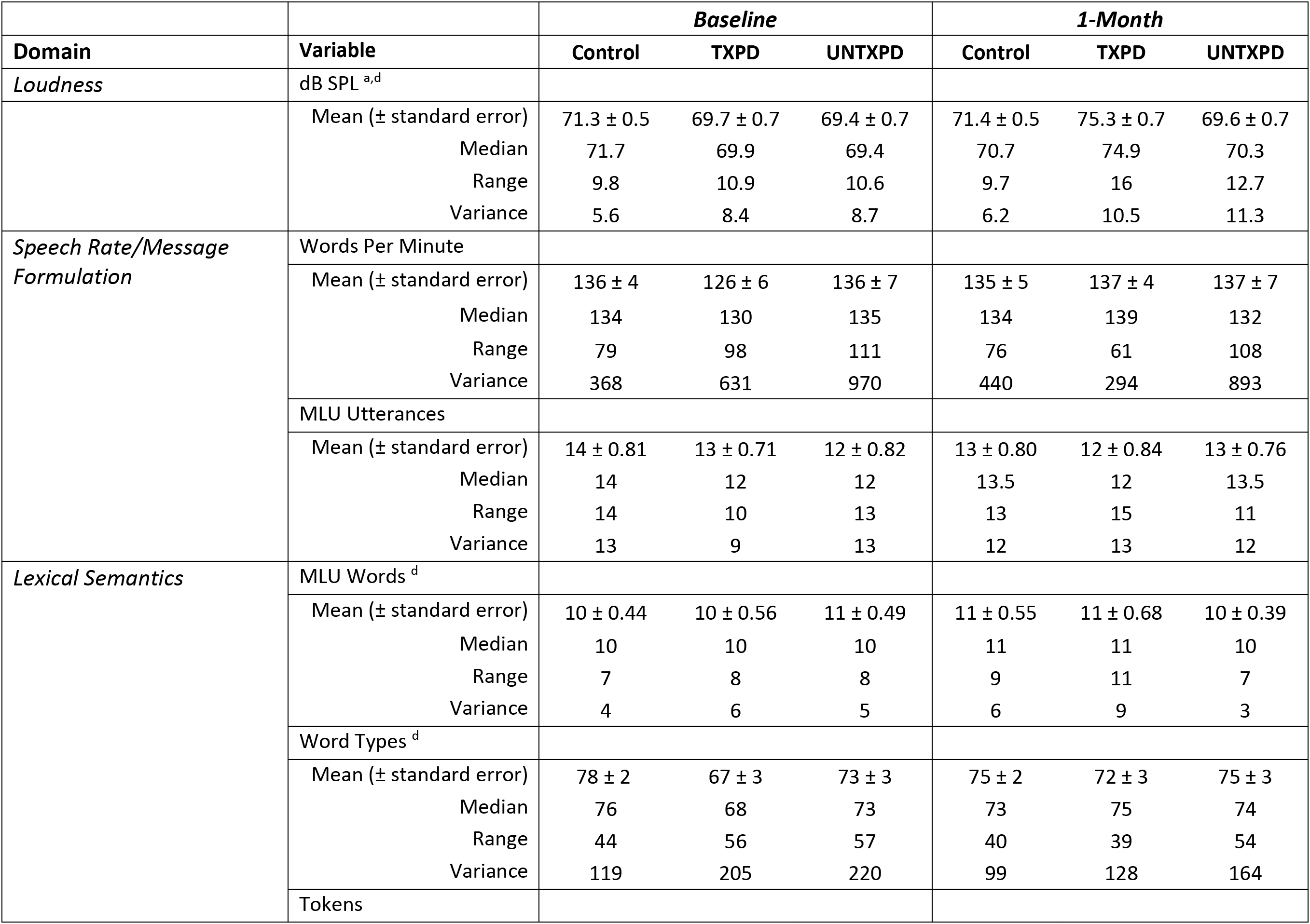
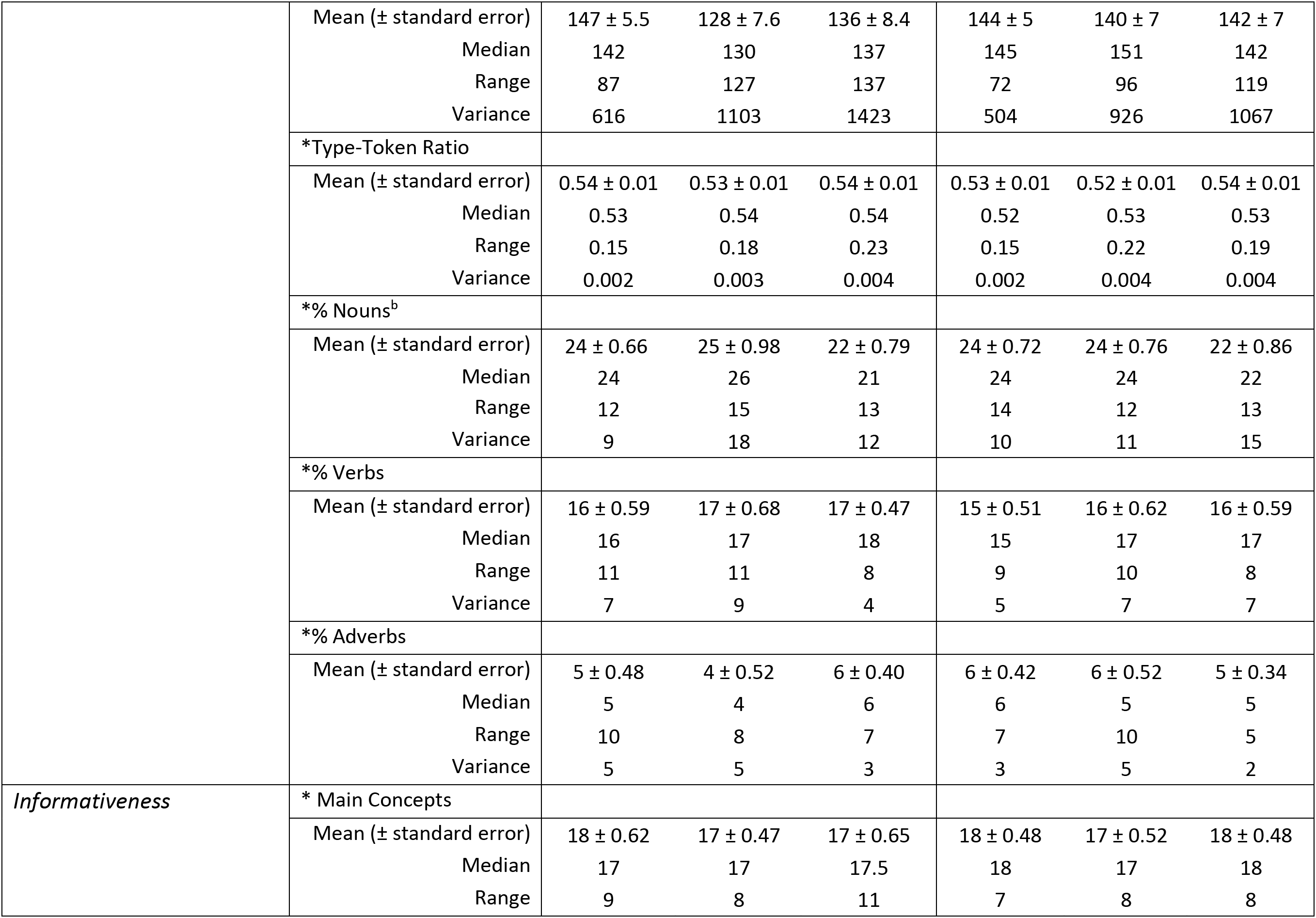
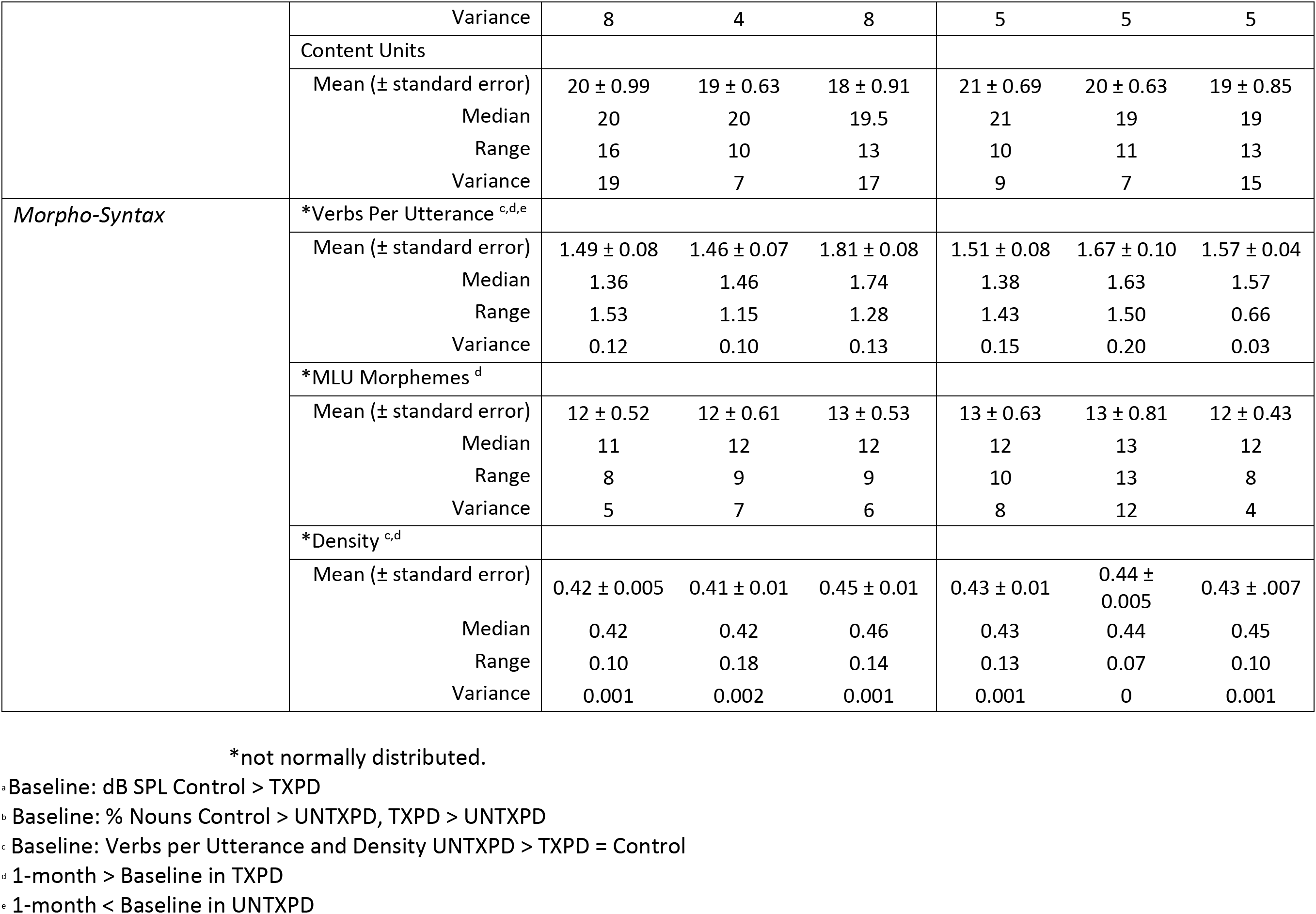
Language variables analyzed for the Cookie Theft picture in Controls, Treated PD (TXPD) and untreated PD (UNTXPD) at baseline and 1-month.

#### Within- and Between-Group Contrasts for Change from baseline to one-month

Group- by-timepoint interactions indicate a change over time that is larger in one group relative to the others – e.g., that performance in the TXPD changed over time to an extent that was significantly greater than that of the other groups. Generalized linear mixed models (GLMM) were conducted in SPSS to account for repeated measures to identify effects of timepoint (baseline, one-month), and to isolate treatment effects in the TXPD relative to the UNTXPD and Control groups. The GLMM was chosen as it combines the linear mixed model, incorporating random effects for variance associated with participants, and the generalized linear models, allowing for analysis of non-normal data using link functions or exponential family distributions (i.e., gamma or inverse Gaussian with log link, Poisson with log link for error counts; (Bolker et al., 2009)). The fixed effects for these models were group, timepoint, and their interaction with the covariance structure set to Diagonal, i.e., with an assumption that variance differed at each timepoint but the correlation between measurements remained constant. The random effect for these models was participant with covariance components structure. Models were estimated with random intercepts and, if model convergence allowed, with random slopes for participants. Effects were considered significant if *p* < .05, Bonferroni corrected for post-hoc multiple pairwise comparisons.

Bivariate Pearson or Spearman correlations, depending on the normality of the distributions, were calculated to identify relationships between change in SPL over time, and language variables demonstrating significant change over time (performance at one-month – baseline). Correlations were considered significant if Pearson *r* or Spearman *rho* > |.50| and *p* < .05, indicating a moderately strong association.

## 3. Results

### 3.1 Participant Demographics

The groups did not differ for age, gender, or handedness distribution, or for *Mini-Mental Status Exam* score (see Table 1 and Supplemental Table 2). However, the PD participants endorsed more depressive symptoms with higher BDI scores than the Controls (TXPD > Controls: contrast estimate = −6.62 [-10.39 - −2.86], *t*(56) = −4.34, *p* < .0001, *d* = 0.86, UNTXPD > Controls: contrast estimate = −4.45 [-7.92 - -0.98], *t*(56) = −4.45, *p* = .009, *d* = 0.58; TXPD = UNTXPD). The groups with PD did not differ for motor symptom severity (Hoehn & Yahr scale, (Bhidayasiri & Tarsy, 2012) or years since diagnosis. One TXPD participant, who was 31 years post diagnosis with PD, was an outlier for disease duration, but was included in the sample since he completed the LSVT LOUD treatment trial and was not an outlier for any of the language variables. Another TXPD participant was an outlier for several language variables, as his picture description was short and simple (i.e., fewer words per minute, MLU words and morphemes, word types, verbs per utterance, density, % verbs [and an extreme outlier for high % nouns], and content units), and thus his data were removed from the study, leaving a total of 19 TXPD participants. As expected, the participants with PD were not as loud as the Control participants when describing the picture (*b* = −5.22 (95% confidence interval = -7.93 - −2.93), *p* < .0001), but the TXPD and UNTXPD groups did not differ from each other for loudness in dB SPL at baseline.

### 3.2. Cross-sectional Group Differences in Language Performance

#### 3.2.1 PD versus Control at Baseline

At baseline, the 49 participants with PD did not differ from Controls for words per minute or for the number of utterances produced during the picture description, nor did they differ for mean length of utterance in words or morphemes, or for lexical diversity (number of different word types or tokens, or for type-token ratio). The groups also did not differ for the informativeness of their descriptions in main concepts or content units. These data are presented in Table 2, and statistical test details are reported in Supplemental Table 2. However, the subset of participants with PD in the TXPD group produced fewer word types than the Controls (contrast estimate = 11.32 [0.68 - 21.96], *t*(56) = 2.63, *p* = 0.03, *d* = 0.34).

The only significant group differences at baseline were for the group of participants with PD randomized to the untreated group (UNTXPD), who produced more verbs per utterance than either the TXPD (contrast estimate = -0.35 [-0.62 - −0.09], *t*(56) = −3.30, *p* = .005, *d* = 0.43) or Control (contrast estimate = −0.32 [-0.57 - 0 0.07], *t*(56) = −3.004, *p* = 0.008, *d* = 0.39) groups (Figure 1). This larger number of verbs per utterance was also reflected in the UNTXPD group producing significantly fewer % nouns and less lexical density than the TXPD group (% nouns: contrast estimate = 3.36 [0.62 - 6.11], *t*(56) = 3.02, *p* = 0.01, *d* = .44; density: contrast estimate = −0.32 [-0.06 - −0.002], *t*(56) = −2.60, *p* = .04, *d* = 0.34) and Controls (% nouns: contrast estimate = 2.74 [0.25 - 5.23], t(56) = 2.54, *p* = 0.03, *d* = 0.33; density: contrast estimate = −0.31 [-0.06 - −0 0.002], *t*(56) = −2.54, *p* = 0.04, *d* = 0.33).

**Figure 1.**
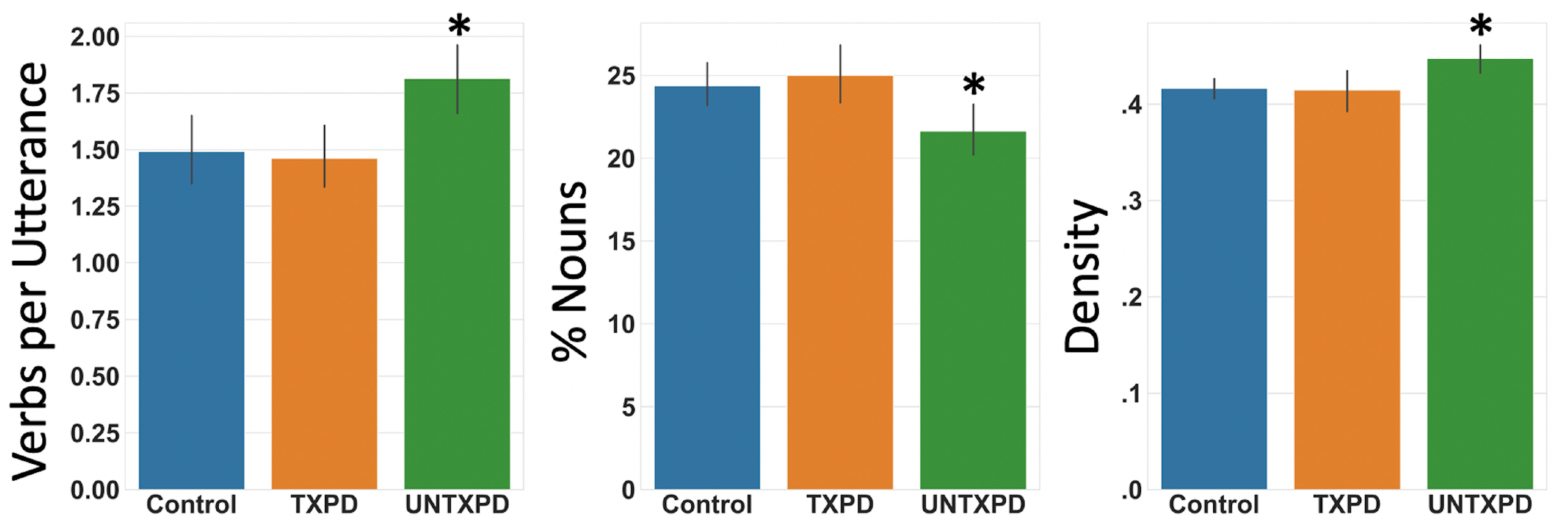
Group differences dependent variables at baseline. The only significant group differences at baseline were attributed to the group of participants with PD randomized to the untreated group (UNTXPD), who produced more verbs per utterance than either the TXPD or Control groups. This higher number of verbs per utterance in the UNTXPD group was also reflected in the percentage of nouns to total words and lexical density (i.e., the number of verbs, adjectives, adverbs, prepositions, and conjunctions to the total number of words). * *p* < .01.

### 3.3 Longitudinal Changes in Language Measures

Group-by-timepoint interactions existed for mean utterance length in words and morphemes, word types, verbs per utterance, and lexical density (parameter estimates and *p*-values reported in Supplemental Table 3, see Figure 2). Post hoc paired contrasts clarified that the TXPD group, with intervening treatment, demonstrated significant increases in the diversity of word types (contrast estimate = 5.37 [1.20 – 9.54], *t*(112) = 1.32, *p* = 0.02, *d* = 0.30), mean utterance length in words (contrast estimate = 1.32 [0.22 – 2.42], *t*(112) = 2.39, *p* = .02, *d* = 0.55) and morphemes (contrast estimate = 1.53 [0.33 - 2.72), *t*(112) = 2.53, *p* = 0.01, *d* = 0.58), and lexical density (contrast estimate = .02 [.004 - .04], *t*(112) = 2.37, *p* = .02, *d* = 0.54) relative to either of the other groups. At the one-month time point, the TXPD group also produced more verbs per utterance (contrast estimate = -0.20 [-0.001 - 0.41], *t*(112) = 1.97, *p* = 0.05, *d* = 0.45) while in contrast, the UNTXPD group produced fewer verbs per utterance (contrast estimate = 0.24 [-0.46 - -0.02], *t*(112) = 2.19, *p* = 0.03, *d* = 0.49).

**Figure 2.**
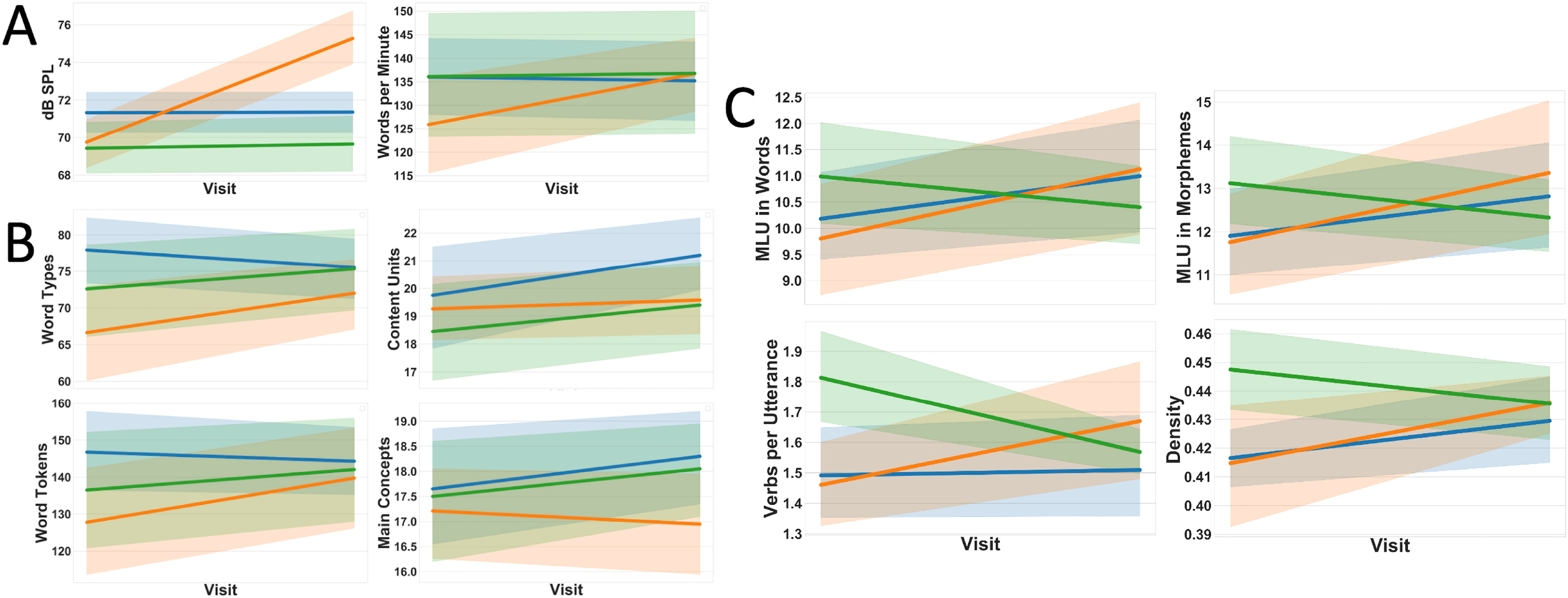
Changes in dependent variables with from baseline to one-month. Significant increases were observed in the TXPD group across speech (A), lexical semantics (B) and morphosyntactic (C) variables assessed in the picture description task. Words per minute (A) and word types (B) increased from baseline to one-month in the TXPD group, but significant group-by-time point interactions were observed only for loudness (A, left) and the language measures indexing morpho-syntax (C) in this study. The interactions indicated that the TXPD group (orange) increased in loudness, words and morphemes per utterance, verbs per utterance, and density given the intervention. In contrast, the UNTXPD group (green) demonstrated a significant decrease in verbs per utterance over the 4-week period between time points. No significant interactions were evident for the language measures indexing lexical semantics (B) in this study. Of note is that the Control group (blue) also demonstrated increases for most of these variables between time points, but the increase was significantly larger in the TXPD group. * p < 0.05, ** p < 0.01, color bands = 95% confidence intervals.

To characterize the increases in utterance length observed in the TXPD group, additional analyses were conducted on the types of words relative to total number of words produced by each participant (e.g., % nouns to total words). Group-by-time point interactions were significant only for % adverbs, with both the TXPD and Control groups producing more adverbs at the one-month time point than at baseline (TXPD: contrast estimate = 1.18 [0.11 – 2.25], *t*(111) = 2.18, *p* = 0.03, *d* = 0.50; Controls: contrast estimate = 1.23 [0.16 – 2.31], t(111) = 2.27, *p* = 0.02, *d* = 0.51).

#### 3.3.1 Change in Language Measures and Change in Loudness

A relevant next question was whether the change observed in language variables in the TXPD group following intervention was related to corresponding change in loudness outcomes (vocal loudness in dB SPL; Figure 3). The TXPD group significantly increased loudness with intervention (TXPD contrast estimate = 5.2 [3.56 – 6.84], *t*(112) = 6.28, *p* < .0001, *d* = 1.44, Supplemental Table 3), while loudness in the other two groups remained fairly constant. The correlations between loudness and language variables at baseline ranged from very small to moderate across groups (Figure 3). In control participants, the only significant correlation indicated that those who were louder at baseline produced more verbs per utterance; all remaining, non-significant correlations were positive (i.e., louder = more) except for % adverbs. In contrast, in the groups of participants with PD, most of the correlations at baseline were negative (i.e., louder = less) except for verbs per utterance in both the TXPD and UNTXPD groups which had positive, but non-significant correlations with loudness (Figure 3).

**Figure 3.**
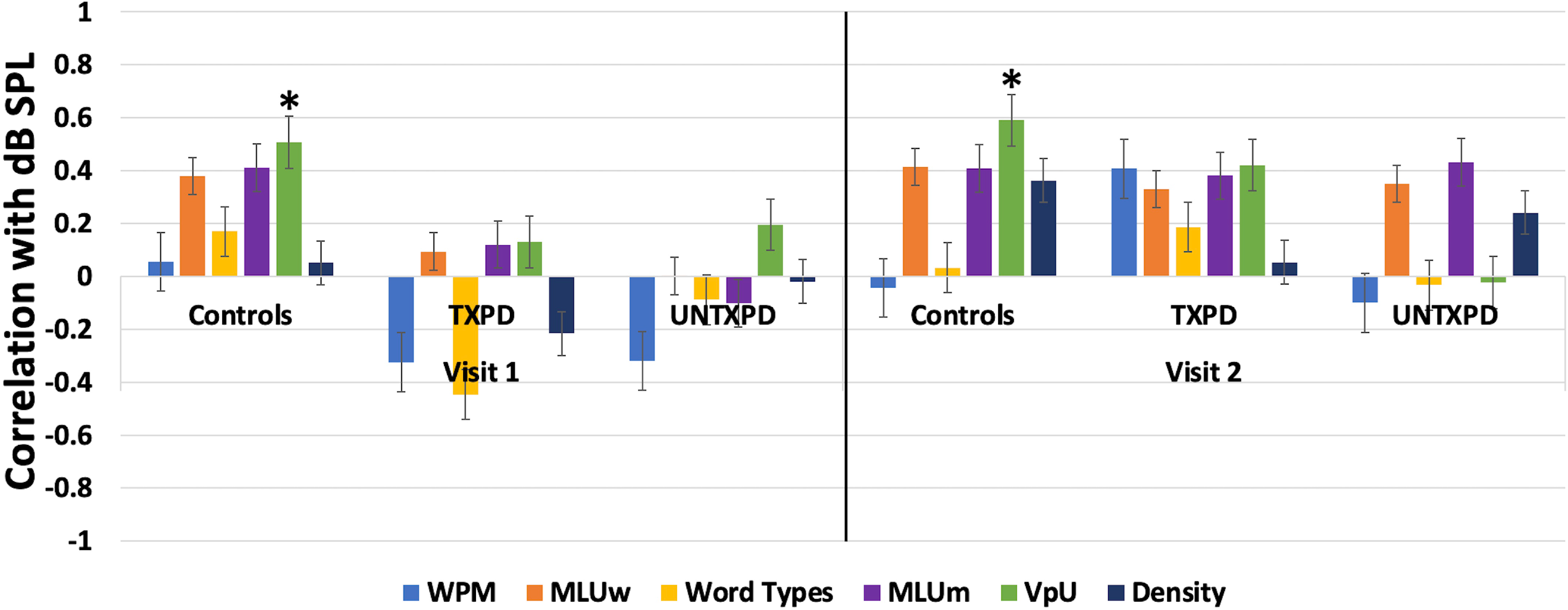
Correlations between loudness and dependent variables and baseline and one-month. Loudness was positively associated with all aspects of language at baseline that were later shown to change following intervention targeting loudness, but only in the Control group. That is, while only the correlation between loudness and verbs per utterance was significant for Controls at baseline, all were positive – i.e., louder = more. Conversely, for both PD groups, loudness was mostly negatively correlated with the language measures – i.e., louder = less. The only positive associations with SPL in the PD groups, though not significant, were for MLUm (TXPD only) and verbs per utterance. The correlations between loudness and language variables ranged from very small to moderate across groups at baseline. The strongest relationships indicated that louder Control participants produced more verbs per utterance. Only weak correlations existed between loudness and language in the PD groups at baseline, which indicated that those who were louder produced fewer words per minute and fewer word types. And, in the TXPD group those who were louder had less density. The latter two points suggest a tradeoff for loudness at the expense of amount and syntactic complexity of verbal expression. At one-month, all participants who were louder produced longer utterances, both for MLU words and morphemes. In the TXPD group, the negative correlation between loudness and words per minute became positive (i.e., louder voice = longer utterances). The relationship between loudness and verbs per utterance became even stronger in the Controls at one-month, increased considerably in the TXPD group (from weak to moderate), but decreased considerably in the UNTXPD group (from weak to none). Finally, the relationship between loudness and density in the TXPD group shifted positively, though it was still a weak correlation; and a similar shift was observed in the UNTXPD. * *r* or *rho* > |.50|, *p* < 0.05.

Relationships at the one-month time point looked quite different. In the TXPD group, all correlations between loudness and language variables were positive except for % adverbs (Figure 3). Although positive correlations with loudness were also evident in the UNTXPD group, particularly for MLU morphemes, these untreated PD participants showed no relationship between loudness and verbs per utterance, demonstrating the inherent variability of language performance on the task.

Perhaps more informative were the correlations between magnitude of change in loudness and magnitude of change in language (Figure 4). Increased loudness, the most direct outcome of the intensive voice treatment, was not significantly correlated with change in any of the language measures in the TXPD group. Rather, while there was only minimal change in loudness in the Control group, increased loudness when describing the picture at the one-month time point significantly positively correlated with words per minute (*rho*(20) = 0.60, *p* = .005). It is important to note that change in loudness was considered “perceptible” (i.e., ≥ 3 dB SPL change, (Fox & Ramig, 1997)) for 16 of the 19 TXPD participants, but only for 3 of the 20 Controls and one of the 20 UNTXPD participants; thus, the range for assessing these correlations in those groups was considerably restricted (see Supplemental Figure 1). The relationships between loudness and words per minute were non-significant in the TXPD and UNTXPD groups and were in opposing directions. The 16 TXPD participants who had gains in loudness had weak and non-significant correlations indicating *reductions* in words per minute (i.e., were slower in describing the picture, *rho*(19) = −0.29, *p* = 0.23) and *more* verbs per utterance (*rho*(19) = 0.30, *p* = 0.21), consistent with previous findings of better use of phrasing given increased loudness (Ramig et al., 1995). In contrast, UNTXPD participants who had minimal increases in loudness (only 1 with a perceptible increase from baseline to one-month) *increased* in words per minute (*rho*(20) = 0.26, *p* = 0.28) as well as in verbs per utterance (*rho*(20) = 0.46, *p* = 0.04).

**Figure 4.**
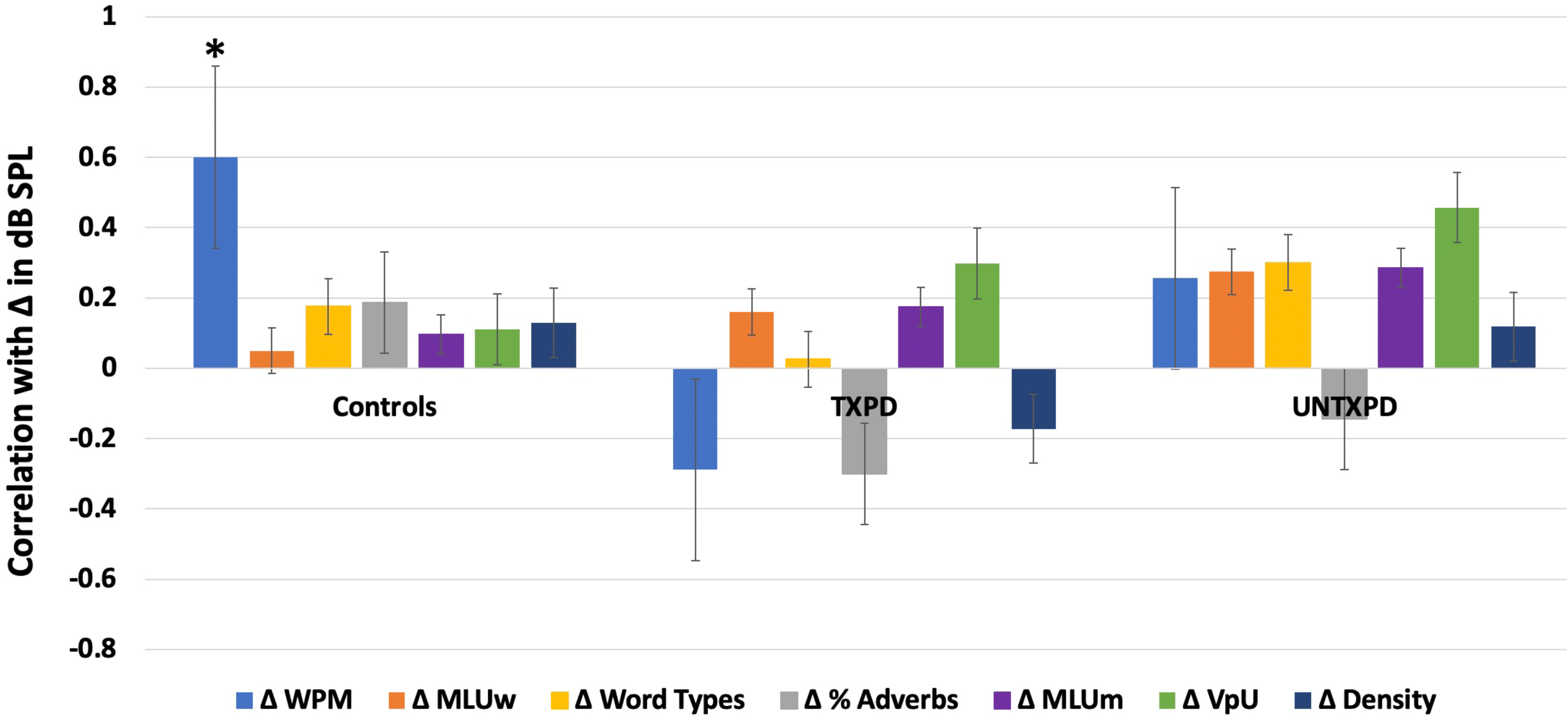
Magnitude of change in loudness and dependent variables by group. Correlations between the magnitude of change (**Δ)** in loudness and the magnitude of change in language measures supported the link between loudness and words per minute, though the relationship was in opposing directions in the TXPD group relative to the Control and UNTXPD groups, supporting the idea that increasing loudness comes at a slight cost for words per minute in the TXPD group, likely associated with the increasing syntactic complexity evidenced by the positive correlation with verbs per utterance. As was evident in Figure 3, the relationship between increasing loudness and longer utterances was consistent across groups. It is important to note, however, that the magnitude of change in loudness was considerably larger in the TXPD group (evident in Figure 5). * *r* or *rho* > |.50|, *p* < 0.05.

## 4. Discussion

Embodied cognition theory, which proposes that language and motor representations of action concepts are tightly intertwined, has been applied to explain impaired action word (verb) processing in PD. Building from this theory, the present study tested whether engagement in an intensive intervention targeting increased vocal loudness might have sufficient impact on the motor system to strengthen action concepts and, in turn, improve retrieval of verbs, or utilization of verbs in more dense syntactic structures in discourse production. Narrative discourse samples elicited from treated and untreated participants with PD, as well as from age-matched controls, in a randomized controlled trial of LSVT LOUD served as a sample of convenience to test whether (1) lexical semantic or morphosyntactic elements of language production were different in PD relative to Controls prior to intervention, and (2) whether treatment-related change in vocal loudness (i.e., motor system) also brought about change in lexical semantics or morphosyntax in language production.

### 4.1 Replication of Group Differences in Language Production

At baseline, the groups generally did not differ for the indices of lexical semantics assessed with the Cookie Theft picture description. They did not differ for lexical diversity (word types, word tokens, or the type-token ratio) or the informativeness of the description content (main concepts or content units). However, relative to morphosyntax, half of the participants with PD produced significantly more verbs per utterance than the other half or the Control group, suggesting considerable within- and between-group variability for verbs per utterance (Table 2). With random assignment of the participants with PD to treatment groups for the RCT, those in the UNTXPD group happened to be those who produced significantly more verbs per utterance compared to the TXPD or Control groups. With the inclusion of more verbs per utterance, the UNTXPD group also produced a lower percentage of nouns and higher lexical density than either of the other groups. These aspects of language production that differed in the UNTXPD group are morphosyntactic elements indicating a difference in the latter stages of processing (Bock & Levelt, 1994; V. S. Ferreira, 2010).

These data do not replicate findings of a verb-specific deficit in PD, at least for language production in the Cookie Theft picture description task. Qualitative characterization of the utterances indicates that the difference in verbs per utterance may relate to the nature of the task rather than to a true difference in verb production. That is, picture description narrative tasks elicit variable performance from individual-to-individual, with some participants simply listing items seen in the picture (i.e., producing no verbs); others producing few utterances but using complete, grammatically correct sentences; and most doing both (“boy, and girl, the boy is falling”). In this cohort, several of the UNTXPD participants (6 of the 20) produced only a few utterances, but all those included verbs, inflating the mean verbs per utterance for this group. Thus, the lack of replication in the present study may be due to the nature of the Cookie Theft picture description task, which differs substantially from other tasks used to elicit verbs in people with PD in the extant literature. Other tasks that have been used for this purpose range from production of verbs in isolation (e.g., verb naming, (Bocanegra et al., 2015)) to lexical decision making or semantic association tasks, which may relate more to recognition or comprehension of verbs than to production (e.g., (Bocanegra et al., 2015; Roberts et al., 2017)). These task-related factors could help explain why the current study did not replicate previous findings of differences in verb retrieval in discourse. Regardless, current findings suggest that the difference in language performance of participants with PD for verb production may not indicate a deficit in lexical semantics.

Beyond task differences, it is also noteworthy that disease duration differed between this and other studies in which differences in verb production were found. For example, Vanhoutte and colleagues (Vanhoutte et al., 2012) compared language production in early (Hoehn and Yahr stages 1-2, mean = 4.7 years duration) relative to advanced (Hoehn and Yahr stages 3-5, mean = 11.4 years duration) disease progression and found that those at advanced stages had more difficulty utilizing action verbs in a story retell task. Similar findings are reported by Bocanegra and colleagues (Bocanegra et al., 2015) who found impaired action verb naming and action semantics in a group of 40 participants with PD (disease duration: 7 years and severity: Hoehn and Yahr stage 2.3), but that only the sub-group of these participants with concomitant mild cognitive impairment also demonstrated impaired syntax and semantics for nouns and action words. Yet, participants with PD, with or without cognitive impairment, did not differ for disease duration or severity, suggesting that the duration of the disease may relate more to the impairment in action word processing, irrespective of the presence of cognitive decline. Thus, again, it is possible that the participants with PD in the present study did not demonstrate specific verb processing deficits because their disease duration was shorter, and they did not have cognitive impairments, at least as indexed by the MMSE.

Aside from verbs specifically, narrative samples from participants with PD tended to include fewer utterances with somewhat reduced lexical density (the number of verbs, adjectives, adverbs, prepositions, and conjunctions to the total number of words) and reduced lexical diversity (i.e., fewer types of words) relative to Controls. These findings are consistent with previous studies involving narratives in PD (Murray, 2000; Murray & Lenz, 2001). That is, participants with PD generally say less and produce less syntactically complex utterances.

The participants with PD in this study also conveyed the same numbers of main concepts and content units as the Controls, thus not replicating differences in the informativeness of narratives in PD (e.g., (McNamara et al., 1992; Murray & Lenz, 2001; Roberts & Post, 2018; Smith et al., 2018)). There are a few apparent differences between the participants in the present study and the other studies reporting differences in content or informativeness in PD. First, the participants with PD in other studies were slightly older than those in the present study, but more importantly they also had longer durations of disease (mean in this study = 5 years, as opposed to 6 years in (Murray & Lenz, 2001), 7.5 years in (Cousins et al., 2018), 8 years in (Vanhoutte et al., 2012), and 9 years in (Roberts & Post, 2018) and (Smith et al., 2018)). The differences in disease duration are important given that there is an increasing potential for cognitive impairment with PD progression. The informativeness of linguistic content production is strongly associated with working memory or processing speed (c.f. Altmann & Troche, 2011), both of which are known to diminish with advancing disease in PD. Thus, the shorter disease duration in current participants, presumably associated with a lower likelihood of mild cognitive impairment, might explain why current participants with PD and Controls did not differ in informativeness.

### 4.2 Did Language Production Improve or Change in the TXPD Group given Intensive Voice Treatment? (i.e., does treating one effector system change another?)

Intensive voice intervention to improve vocal loudness was associated with increased speech rate (words per minute) and mean length of utterance in the TXPD group. Because the intervention focuses on the respiratory-laryngeal-oromotor target of vocal loudness, it is likely that the increase in respiratory support corresponded not only with increased loudness, but also with increased breath support enabling production of longer utterances. However, the TXPD group also had significant increases in the number of morphemes per utterance (i.e., morphological markers like plural -s, determiners, auxiliary verbs, etc.). This suggests not only an increase in utterance length, but also in morphosyntactic complexity in the TXPD group. The latter is further supported by the finding of increased lexical density in the TXPD group, where density is indexed by the number of verbs, adjectives, adverbs, prepositions, and conjunctions to the total words produced. Thus, the TXPD group produced more words and more syntactically complex utterances after intervention. In fact, at the one-month time point, the TXPD group no longer differed from the Controls for any language variable.

There also was a significant increase in the numbers of verbs per utterance in the TXPD group and a significant decrease in verbs per utterance in the UNTXPD group between time points. As mentioned above, there was considerable variability in verbs per utterance at baseline which increased in the TXPD but decreased in the UNTXPD at the one-month time point. Again, the nature of the task and the tendency to list items and not produce complete utterances, as discussed above, may have contributed to this. However, verbs per utterance significantly correlated with loudness at both assessment time points in the Control group, and only at the one-month assessment in the TXPD group after completion of the intervention. That correlation was weak at baseline and non-existent at the one-month time point in the UNTXPD group. The existence of a relationship between loudness and verbs per utterance is potential support for the study hypothesis that treating one effector system might influence the other, but this relationship did not hold when looking at the correlation of the magnitude of change in loudness to the magnitude of increase in verbs per utterance in the TXPD group. That is, there was not a larger increase for verbs per utterance in the TXPD participants with a larger increase in loudness.

While overall the group data demonstrated increases in morpho-syntactic complexity with improved loudness in the TXPD group, those with the biggest boost in loudness had a *reduction* in their production of words per minute and lexical density. Magnitude of change in loudness significantly correlated with the magnitude of change in rate of speech, or words per minute, only in the Control group in whom the change in loudness was non perceptible. The same pattern of positive correlation between change in loudness and words per minute was seen in the UNTXPD group, but the opposite was seen in TXPD participants (Figure 4). Similar but non-significant negative relationships were observed for TXPD participants with greater increases in loudness associated with slight decreases in lexical density in the TXPD group, whereas opposing effects were observed in the Control and UNTXPD groups.

The latter findings, of slower or less complex language in participants with larger gains in loudness with intervention, suggest a subtle tradeoff for loudness in exchange for the resources required for syntactic complexity of verbal expression. The attentional focus on loudness that is central to the intervention, and to the context in which the assessments took place, may require cognitive resources that are thus limited for the language demand of the task. This is not an impairment of language, but rather an indication of the realities of the constraints of resource capacity (c.f., Kahneman, 1973). As well, the use of better respiratory-laryngeal support may have allowed for better phrasing of utterances, requiring slightly slower speech rate. Further study of these participants with PD in the maintenance phases of recovery (e.g., 3 or 6 months after TX ends) might elucidate whether the tradeoff lessens when the “new normal” of speaking loudly has gained automaticity.

It is known that the efficiency of allocating attention during language performance while doing other tasks (e.g., swallowing (Troche et al., 2014), walking (Rochester et al., 2014)) is altered (e.g., slower production of sentences with increasing syntactic complexity by children who stutter (Usler et al., 2017)). When there is a conflict between the resources needed for speech motor control and for language processing, both tend to become more variable (e.g., (MacPherson & Smith, 2013)), and even healthy adults tend to sacrifice speech rate to preserve syntax and content (Kemper et al., 2011). In participants with PD, loudness becomes more variable during a spontaneous discourse when there is a greater cognitive demand (i.e., extemporaneous speaking) relative to a more automatic one (i.e., reading) under dual-task conditions (Whitfield et al., 2019). While the speaking task in the present study did not have a dual-task condition, it may be that the attentional focus on loudness, which successfully boosted loudness at the one-month time point in the TXPD group, came at a cost for the later processing stages of discourse production. In fact, at the one-month time point, within group variance was larger for the TXPD group for all morphosyntactic metrics (MLU in words and morphemes, verbs per utterance) as well as for a metric of discourse planning (i.e., number of utterances, main concepts), but was less variable for all these measures in the UNTXPD group. Investigating within-participant variability at each time point (e.g., at 3- and 6-month follow up sessions) would provide more evidence for or against this proposed explanation.

To our knowledge, there is only one previous study that investigated change in language production in participants with PD following a motor-based behavioral intervention. Altmann and colleagues (Altmann et al., 2016) compared sentence-level picture description performance (describing a single action depicted in a picture in one sentence without the use of pronouns) in participants with PD before and after either an aerobic exercise program or a stretch-balance program. The sentences were analyzed for completeness (included characters and verb), fluency (pauses or false starts), and grammatical accuracy. These authors hypothesized that aerobic training, which had been shown previously to improve cognition and memory, would improve cognitive and executive function, and in turn improve language. They found that aerobic training in the PD group improved completeness, but there were no significant changes in fluency or grammaticality. That study did not assess change in motor system function to determine whether one of the training types was more or less likely to improve motor function. Nonetheless, these findings are like the data reported here, i.e., that verb use in utterances/sentences improved with motor training. Altmann and colleagues did not carefully assess morpho-syntax in their participant’s responses, instead assessing for correctness of grammatical structure.

There are limitations to the study that preclude attributing the changes in language production in the TXPD group conclusively to the motor system involved in improved vocal performance. First, the utility of a hierarchy of speech tasks for practicing the loud voice in context not only provides vocal practice, but also extends practice to language production at both lexical semantic (e.g., word association, word list generation, and naming tasks) and morpho-syntactic levels (e.g., practicing loudness while reading sentences and paragraphs, generating longer utterances in spontaneous conversation, and practicing salient communication activities such as making phone calls or giving lectures/presentations). While the focus of this practice is on “speaking loud”, the exposure to daily language processing may be as responsible for the observed changes in language use as the vocal practice. Further, the UNTXPD and Control groups did not receive similar language exposure during the four-week period between assessments. Thus, future study is warranted, perhaps comparing the language performance in participants with PD receiving LSVT LOUD and PD participants receiving LSVT ARTIC (focus is on pronunciation rather than loudness, but with similar hierarchical speech tasks), as in Ramig et al. (2018), or LSVT BIG with no focus on voice/speech production.

## 5. Conclusions

The central hypothesis of the study is that improving function in the motor system might influence the representations of action words, aligning with the ideas of embodied cognition. Individuals with PD who underwent an intensive voice treatment to increase vocal loudness had concomitant increases in the later stages of language production, or morpho-syntax, during a picture description discourse task. More specifically, increased verbs per utterance in the TXPD group aligns with previous literature positing that the primary difference in language production in PD centers on action word processing (verbs). Given that the verb is the core of sentence production with morphology and syntax falling in line after the verb is selected, it is likely that the increase in verbs per utterance led to use of more morpho-syntactically complex sentence structures - i.e., more lexically dense utterances. Future work can indicate whether the verbs that increased in this sample were action verbs, or other types of verbs (state, cognitive), which would lend more support to the embodied cognition literature and the link between the motor and language systems.

## Supporting information

Ramage_Supplemental Materials

Ramage_Supplemental Table 2

## Data Availability

Data are held by the authors and may be available upon request.

## Acknowledgements

We are grateful to the participants in this study who provide us inspiration to continue working to improve speech and language in the PD population. The LSVT LOUD treatment trial was funded by grant R01DC0115 from the National Institute on Deafness and Other Communication Disorders (Principal Investigator: Ramig). We also thank Kate Phelps and Shannon Bryant for their help in transcribing and verifying transcript accuracy.

## Notes

### Competing Interest Statement

The authors have declared no competing interest.

### Clinical Trial

NCT00123084

### Author Declarations

data were collected at the National Center for Voice and Speech-Denver, an affiliate of the University of Colorado-Boulder (UCB). Additional screening/inclusion and demographic data were collected from neurology and otolaryngology offices in Denver, and the radiology department of the University of Colorado Health Sciences Center-Denver (UCHSC). Study procedures were approved by the institutional review board at the University of Colorado at Boulder and are shared at ClinicalTrials.gov identifier: NCT00123084. All participants provided informed written consent for participation.

