## Supplementary material for "Improved morpho-syntax in discourse following intensive voice treatment in Parkinson’s disease: Secondary outcome variables from a Randomized Controlled Trial (RCT)": Ramage_Supplemental Materials

| **Block** | **ICC(2,1)>.90 (Excellent)** | **ICC(2,1)= .75-.90 (Good)** | **ICC(2,1)<.75 (Moderate)** |
| --- | --- | --- | --- |
| **1** | TTR, total utterances, words, %nouns, %prepositions | Density, verbs per utterance, %adjective | %verb, %adverb |
| **2** | TTR, total utterances, words, %nouns | Density, %verbs, %prepositions, %adjectives | Verbs per utterance, %adverbs |
| **3** | TTR, density, verbs per utterance, total utterances, words, %verbs, %nouns, %prepositions, %adjectives |  | %adverbs |
| **4** | TTR, total utterances, words, %verbs, %nouns, %prepositions | Verbs per utterance, %adjective, %adverb | Density |
| **Total** | TTR, total utterances, words, %nouns, %prepositions | Density, verbs per utterance, %verbs, % adjective, %adverbs |  |

**Supplemental Table 1**. Interrater reliability, using intra-class coefficient (ICC), for EVAL.

Supplemental Tables 2 and 3 are .xlsx spreadsheets.

**Supplemental Figure 1**. Scatter plots demonstrate the magnitude of change in SPL in the TXPD group relative to the other two groups, and its relationship to change in words per minute and verbs per utterance.

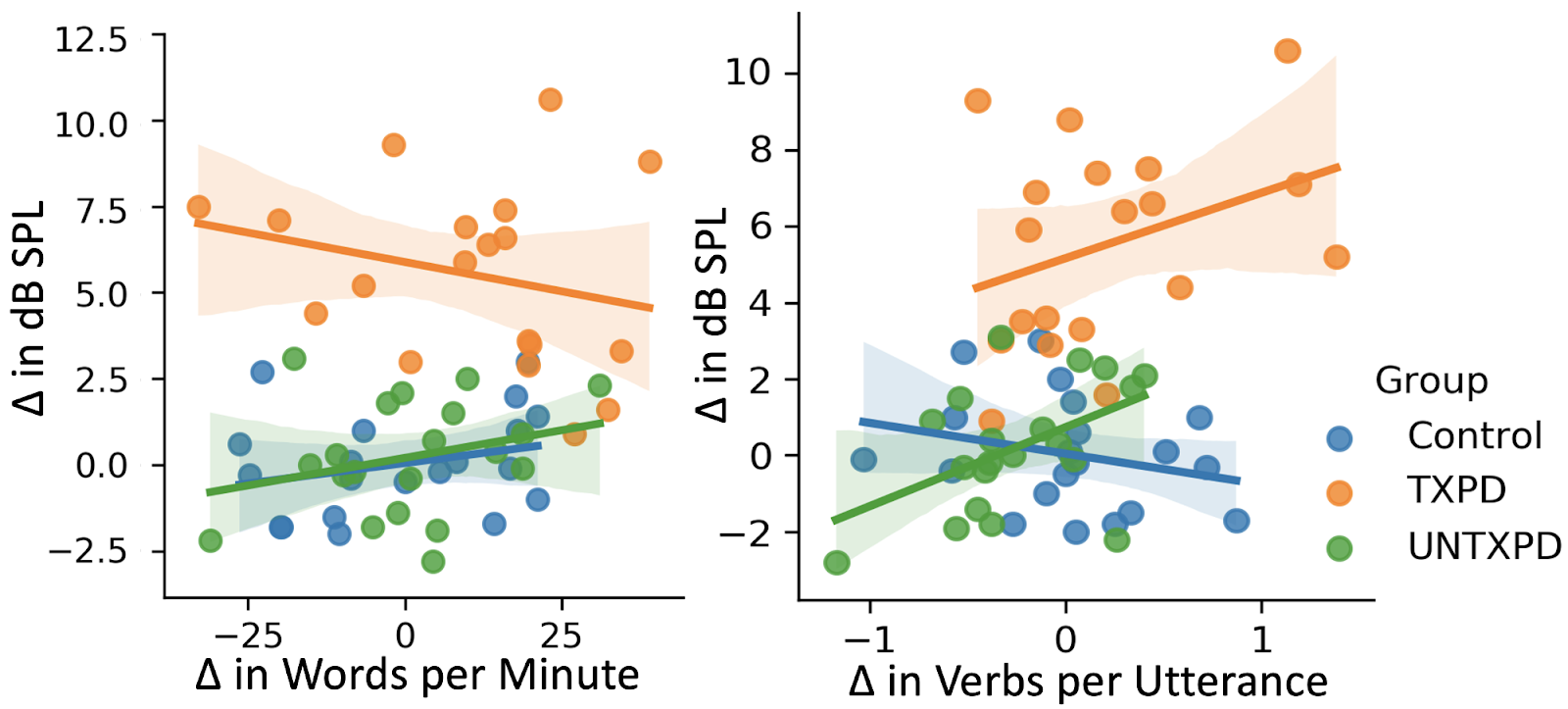
